# Fungal infections in Sudan: an underestimated health problem

**DOI:** 10.1101/2023.06.20.23291647

**Authors:** Sarah A. Ahmed, Mawahib Ismail, Mohamed Albirair, Abdelsalam Mohamed Ahmed Nail, David W. Denning

## Abstract

Fungal diseases are associated with high morbidity and mortality, yet their epidemiology and burden are not well addressed. While deaths probably exceed 1.5 million per year, many cases remain undiagnosed and underreported. Estimating the burden of these diseases is needed for prioritization and implementation of effective control programs. Here we used a model based on population at risk to estimate the burden of serious fungal infections in Sudan. The prevalence of the susceptible population including HIV, TB, cancer, asthma, and COPD was obtained from the literature. Incidence and prevalence of fungal infections were calculated using local data when applicable and if not available then regional or international figures were used. In total, the estimated number of Sudanese suffering from fungal disease is 5 M (10% of the total population). Tinea capitis, recurrent vulvovaginitis and keratitis are estimated to affect 4,127,760, 631,261, and 6,552 patients, respectively. HIV-related mycosis is estimated to affect 5,945 oral candidiasis, 1,921 esophageal candidiasis, 571 *Pneumocystis* pneumonia, and 462 cryptococcal meningitis cases. *Aspergillus* infections are estimated as follow: 3,438 invasive aspergillosis, 14,950 chronic pulmonary aspergillosis, 67,860 allergic bronchopulmonary aspergillosis cases, while the prevalence of severe asthma with fungal sensitization and fungal rhinosinusitis was 86,860 and 93,600 cases, respectively. The neglected tropical disease eumycetoma was estimated to affect 16,837 cases with a rate of 36/100,000. Serious fungal infections are quite common in Sudan and require urgent attention to improve diagnosis, promote treatment, and develop surveillance programs.

**Author Summary:** Fungal infections are present globally, but the burden is higher in sub-Saharan Africa. Many of these infections occur because of underline clinical conditions such as HIV, TB, cancer, asthma, and chronic obstructive pulmonary disease (COPD), while some may occur without underline conditions. We used the prevalence of susceptible populations to estimate the burden of fungal diseases in Sudan. Based on that, we found a total of 5 million Sudanese suffering from fungal infection. The most common disease was scalp infection (tinea capitis), followed by recurrent vulvovaginitis. Fungal eye infection was estimated to affect 6,552 patients, and the subcutaneous neglected tropical disease eumycetoma affects 16,837 individuals. We also reported a high burden for HIV- and TB-related mycoses. The study highlighted the need to develop surveillance programs for fungal infections in Sudan.

## Introduction

The African continent has the highest prevalence of infectious diseases in the world leading to many years of life lost (YLL) and high disability-adjusted life years (DALYs) lived [1]. Estimated deaths as a result of communicable diseases in 2019 was 4.1 million. The high death rate is mainly attributed to human immunodeficiency virus (HIV), respiratory infections, malaria and tuberculosis (TB) among others [2]. Malnutrition, poverty, and illiteracy are the main drivers, as 34 out of 54 African countries are classified as low or lower middle income countries [3]. Of the total global HIV and TB cases, 75% and 25% are found in Sub-Saharan Africa, respectively [4, 5]. With the introduction of antiretroviral therapy and national TB programs in many countries, the control of these infections has remarkably improved. However, mortality remains high mainly because of other comorbid conditions such as fungal infections [6]. Around 50% of opportunistic infections affecting HIV individuals are thought to be caused by fungi. Cryptococcal meningitis and *Pneumocystis* pneumonia are the most common AIDS-related mycotic diseases with an annual reported incidence of 220,000 and 400,000 respectively [7, 8]. Patients with pulmonary TB are at moderate risk for the development of chronic pulmonary aspergillosis (CPA), with an estimated prevalence of ∼3 million cases [9]. Notably, a large proportion of these infections occur in Africa [10]. Despite the high burden of fungal diseases in Africa and worldwide, they have received little attention by healthcare organizations. That is mainly because mycotic infections are challenging in terms of diagnosis, treatment and control [11].

Fungal infections in humans are mainly caused by about 30 species of fungi, with a large diversity of other species as occasional pathogens or allergens. Infections range from superficial affecting skin, hair, and nail to deep systemic and invasive conditions [12]. Invasive infections are generally caused by a few species and occur mainly in patients with underlying diseases or under immunosuppressive medications [11]. In such patients, the most commonly reported fungi are *Candida*, *Pneumocystis jirovecii*, *Cryptococcus*, and *Aspergillus*, with other important potentially lethal opportunists including *Mucorales*, *Histoplasma, Talaromyces,* and *Emergomyces* [12]. Patients with cystic fibrosis, chronic lung diseases or anatomically impaired lungs are at risk of infection by many fungal species, mainly *Aspergillus*. Serious cutaneous or subcutaneous infections include sporotrichosis, chromoblastomycosis, mycetoma, and phaeohyphomycosis [12].

In recent years, efforts have been made by Global Action for Fungal Infections (GAFFI, https://gaffi.org/) and the Leading International Fungal Education (LIFE, https://www.fungaleducation.org/) to estimate the burden of fungal diseases both globally and in many countries in the world. In Africa, and for the first time, the burden of mycotic diseases has been studied and estimated for about 23 countries [13]. In all these studies, reliable data on fungal infections are limited or unavailable, particularly for entities other than cryptococcosis. In addition, the estimated incidence and prevalence of fungal diseases was found to be much higher than previously assumed or reported. For example, studies have shown that, around 8% of the Ethiopian population and 11.5% of Eritrean population suffer annually from serious fungal infections [14, 15]. Of these infections, cryptococcal meningitis, *Pneumocystis* pneumonia and aspergillosis are the most frequent potentially lethal conditions. However, reported cases attributed to these fungi in countries like Ethiopia or Eritrea are few.

The Republic of Sudan, located in Northeast Africa, is bordered by Egypt, Eritrea, Ethiopia, South Sudan, Central African Republic, Chad and Libya. With a total area of 1,882,000 km^2^, the country is the third largest country in Africa. It has been independent since 1956 and to date, Sudan has suffered internal conflict, civil war, and political instability. More than half of the population resides in rural areas and around 46% live below the line of poverty [16]. Sudan is endemic with mycetoma with more than 7,000 cases reported by the Mycetoma Research Center (MRC) in Khartoum [17]. Despite that, mycetoma has been badly neglected for many decades and only after the advocacy of MRC and mycetoma consortium was the disease has been added to the World Health Organization (WHO) list of Neglected Tropical Diseases (NTDs) along with chromoblastomycosis and sporotrichosis [18]. In the present study, we aimed to investigate the burden of serious fungal diseases in Sudan and provide a clear view of the mycotic diseases profile in the country.

## Methods

The basis of our estimates is the use of appropriate denominators of at-risk populations and applying numerators for different fungal diseases. We sourced total population and the number of children (<15 years) from the CIA fact book [19]. To estimate chronic obstructive pulmonary disease (COPD), we used the ratio of those over 40 years to the total from UN estimates for 2020 [20]. We took data about HIV prevalence, proportion on antiretroviral therapy (ART), and AIDS-related deaths from UNAIDS for 2020 [21]. We calculated the proportion at risk of opportunistic fungal infections by assuming a 7-year decline in CD4 counts to <200x 10^6^ combined with an average of 11% failing ART [22]. Pulmonary TB was taken from the WHO Global tuberculosis report 2020 [23]. We assumed a 10% rate of asthma in adults, taken from the 2003 World Health Survey in Ethiopia [24]. Unpublished data indicates a 16% prevalence of COPD in those over 40 years [25] and we have assumed a 10.5% admission to hospital (or equivalent healthcare contact) for exacerbations. Lung cancer and acute leukaemia for 2020 were taken from the International Agency for Research on Cancer [26] and the acute myeloid leukaemia rate from the WHO estimates for low-income countries (2.5/100,000) [27]. Annual renal transplantation procedures for 2020 (possibly lowered by COVID-19) were taken from the International Registry on Organ Donation and Transplantation [28].

For the prevalence and incidence of fungal disease in Sudan, we searched PubMed, Web of Science and Google Scholar databases. We also searched scientific repositories and health reports published by national and international professional organizations. The initial search was performed using the term “Sudan” and “fungal infection” or “Sudan” and “mycosis”. We searched the databases for individual fungal disease and underline risk groups. If no data was available, we calculated the burden using regional or international figures.

## Results

In 2021, the estimated population of Sudan was 46.8 million; of them 42% were children and 20% were adults over 40 years [19] (Table 1). According to the World Bank data, Gross Domestic Product (GDP) per capita in Sudan is around 764.3 US$ [29]. The public health infrastructure in the country is underdeveloped with a very low health expenditure of only 1.15 % GDP. There are only 1.6 primary health care facilities and 6.7 hospital beds per 10,000 population. Low wages and income instability have resulted in high attrition of Sudanese medical staff leaving only 4.1 physicians and 8.3 nurses per 10,000 inhabitants [30, 31].

**Table 1.** Demographic data and the population at risk for fungal infections in Sudan.

| Data | Number | Comment | Reference |
| --- | --- | --- | --- |
| Population | 46.8 million | 2021 | [19] |
| Children 0-14 | 19.7 million | 42% of the population | [19] |
| Adults over 40 years | 9.36 million | 20% of the population | [20] |
| HIV/infection | 49,000 | Prevalence of HIV/AIDS 2020 | [21] |
| Pulmonary TB | 28,000 | Annual incidence 2020 | [23] |
| Asthma in adults | 10% | Taken from Ethiopia (2003) | [24] |
| COPD patients | 1,360,800 –<br>2,214,950 | Population, 3.5%, extrapolated from Egypt, 16% of<br>>40 years unpublished data from Sudan | [25, 32] |
| COPD hospital admissions | 142,900 – 232,570 | 10.5% annually, extrapolated from Algeria | [33] |
| Lung cancer | 647 | 1.4/100 000 incidence 2020 | [26] |
| Acute leukaemia | 1,297 | 2.7/100,000 incidence | [27] |
| Renal transplantation | 139 | 2020 | [28] |

Among the sub-Saharan African countries, Sudan is considered to be one of the lowest HIV-burden countries with an estimated prevalence among adults of 0.1% in 2020. Of the estimated 49,000 people living with HIV (PLHIV) in 2020 only 18,000 knew their status and 12,000 were on antiretroviral therapy, while the estimated death is about 2,300 [21]. Although invasive fungal diseases on the global scale contribute to about 50% of AIDS-related death annually [22], no available data on fungal disease related death in HIV patients in Sudan. Therefore, we estimated the burden based on figures from other studies [8, 34, 35]. We estimated the annual incidence of AIDS related fungal diseases in Sudan to be: 462 cases of cryptococcal meningitis, 571 cases of *Pneumocystis* pneumonia, 1,921 of oesophageal candidiasis, and 5,945 of oral candidiasis. Although histoplasmosis is an AIDS defining disease and has been reported to coexist with TB/HIV in sub-Saharan countries [36], we could only find two case reports [37, 38].

In addition to HIV, respiratory diseases such as COPD and cancer are among the risk factors for the development of invasive aspergillosis [39]. The prevalence of COPD in Sudan was found to be 16% in those over 40 years of age, and the annual incidence of lung cancer and acute leukemia was 647 and 1,297 cases, respectively [26, 27]. We assumed that 13% of cases of acute myeloid leukaemia develop invasive aspergillosis [40], and an equal number of cases in other leukaemias and lymphomas [35, 40], also that 4% of HIV deaths are linked to invasive aspergillosis [41]. Based on this data, we have estimated the overall annual incidence of invasive aspergillosis to be 3,438 cases (35 per 100,000).

Closely linked to malnutrition and poverty, TB remains one of the major health issues in Sudan with an infection rate of 63 per 100,000 population and a mortality rate of 9.81 per 100,000 population [23]. Some patients with negative tests for TB are included in these figures, and some will have alternative diagnoses such as chronic pulmonary aspergillosis (CPA). After treatment for pulmonary TB, lung cavitation develops in 20% to 40% of patients. This leads to colonization by *Aspergillus* species and in some the subsequent development of CPA [42]. We estimated CPA following TB infection in Sudan using the recently published model for India, and data from Africa [43, 44]. CPA cases which are mistaken for, are co-infections with or occur in the months after therapy for TB are estimated at 3,359 cases annually, of whom around 359 die, accounting for 11.7% of TB all deaths. The cumulative prevalence of CPA including those cases and those occurring post TB cure over 5 years is estimated at 14,950 cases.

A community-based survey on asthma conducted in 4 cities in Sudan estimated the prevalence of the disease in adults to be around 10% [45]. A slightly higher prevalence of 12.5% has been reported in children [46], and under-diagnosis is very common [47]. Allergic bronchopulmonary aspergillosis (ABPA) occurs mostly as a complication of asthma and other diseases affecting respiratory function such as cystic fibrosis (CF) [48]. In Sudan, cystic fibrosis is relatively rare and only 35 cases have been reported [49]. The 2,714,400 adults living with asthma in Sudan are probably at risk of ABPA, and to estimate the burden we assume that 2.5% develop ABPA according to studies conducted in South Africa, Saudi Arabia and Uganda [50–52]. The likely prevalence of ABPA in Sudan can then be calculated as 67,860 (145 per 100,000). The severity of asthma usually increases in those with sensitization by *Aspergillus* or other fungi and has been reported in 42% of Ugandan adults with asthma [52]. To estimate the burden of severe asthma with fungal sensitization (SAFS), we assume that severe asthma occurs in 20% of the total asthmatic population and of those, 16% have SAFS. Thus, the prevalence of SAFA in Sudan is conservatively estimated at 86,860 cases (191 per 100,000). There may be some overlap between ABPA and SAFS, but patients with allergic bronchopulmonary mycosis [52] have been omitted from the estimates.

Depending on the infected host, the clinical presentation of fungal rhinosinusitis can range from allergic to granulomatous or invasive and life threatening. The prevalence of the allergic rhinosinusitis ranges from 10% to 40% and of these 5% are assumed to be caused by fungi and we have used a 2% rate [53, 54]. Accordingly, we estimated that allergic and granulomatous rhinosinusitis in Sudan possibly affect as many as 93,600 people, a prevalence of 200 per 100,000. Veress et al. [55] studied 46 cases of granulomatous sinusitis from Sudan due to *Aspergillus*. Additionally, in around 4 years, Ahmed and Osman [56] reported their experience with 440 cases of fungal rhinosinusitis from two centers in Khartoum. Furthermore, of the 579 nasal biopsy samples studied by Mahgoub et al. [57], 280 were positive for fungi, and at least eight patients were diagnosed with chronic invasive fungal sinusitis [57].

Candidemia and *Candida* peritonitis are particularly serious infections affecting critically ill patients, such those in ICU. We couldn’t find data on these infections in Sudan. Internationally, the incidence of candidemia has been estimated to occur in 2 to 21 per 100,000 [10]. We have applied a low international figure (5 per 100,000) to determine the annual incidence in Sudan, a total of 2,340 cases. *Candida* peritonitis in surgical patients is estimated to infect 351 cases per year if we assume that 33% of candidemia cases are ICU patients and 50% of them might develop *Candida* peritonitis [58].

Globally, around 138 million women are affected yearly by recurrent vulvovaginal candidiasis (rVVC), occurring at least 4 episodes per year, and the prevalence among African women is 33% [59]. In Sudan, Kafi et al. [60] reported a prevalence of 10.1%, while in pregnant women who are at higher risk, the reported prevalence was 16.6% and 32.6% according to Nemery et al. [61] and Abdelaziz et al. [62], respectively. In general, women in the age bracket of 15-50 years are at risk of rVVC. Using a conservative 6% proportion on this age group [63], around 631,300 Sudanese women are probably affected by rVVC (2698 per 100,000).

Cases of entomophthoramycosis were reported from Sudan: 10 cases of basidiobolomycosis and one caused by *Conidiobolus* spp. [64–67]. The latter case was an immigrant from Sudan who was diagnosed one month after arrival in Switzerland. For mucormycosis, we estimated an annual incidence of 94 cases with an infection rate of 0.2 per 100,000 population, based on data from France [68].

Sudan is an endemic region for mycetoma; the incidence in a single endemic village was found to be 14.5 per 1,000 inhabitants [69]. This long-standing and disabling disease carries a significant burden to each patient and their community. About 86% (5,819) of the mycetoma patients reported by the MRC had have the infection for nearly 10 years. Yet, the exact prevalence of the disease in Sudan is unknown. Abbott reported a total of 1,231 patients admitted to Sudanese hospitals in a period of 2.5 years [70]. Using a geostatistical model, Hassan et al. [71] estimated the burden of the disease over 28 years to be 51,541 cases. Based on that, we calculate a total of 1,841 new cases annually. Assuming a 2% annual mortality and an infection duration of 10 years, we estimate an annual total of 16,837 prevalent cases, with a prevalence of 36 per 100,000. For other implantation mycosis: we did not find any report for chromoblastomycosis and only found two cases of sporotrichosis in the literature [72].

Tinea capitis was reported in 3.22% of school children in Sudan [73]. More recent data from the Eastern part of the country showed a prevalence of 17% [74]. We estimated the prevalence from the total number of children who are risk of infection using 19%, and probably 4,127,760 are affected [75].

For fungal keratitis, we could not find epidemiologic data from Sudan. Therefore, we estimated the burden based on the data from Egypt as 14 per 100,000 individuals which gave an annual incidence of 6,550 patients [76].

## Discussion

Like other African developing countries, Sudan suffers from an excessive burden of both communicable and non-communicable diseases [30]. In the present study, and for the first time, the burden of serious fungal infections in Sudan is investigated. According to our results, around 5 million Sudanese are suffering from fungal diseases; representing 10% of the total population (Table 2). Though very high, the burden is similar to that reported from Ethiopia, but higher than Egypt [14, 76]. That is mainly because these estimates were based on the population at risk, and therefore variability between countries is anticipated. Furthermore, Sudan and Ethiopia are low-income countries sharing similar economic conditions while Egypt is lower middle-income country.

**Table 2.**
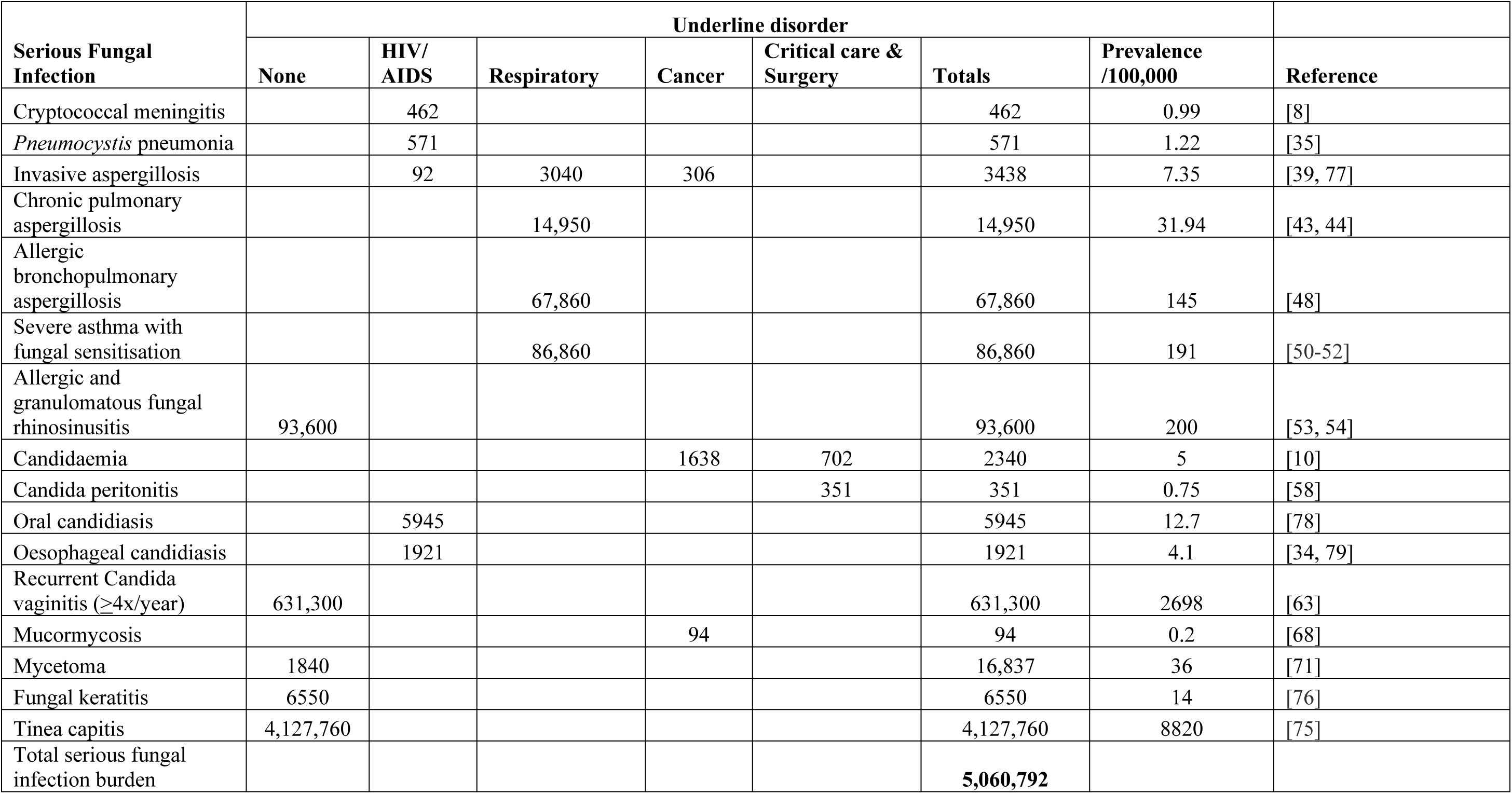
Estimated burden of serious mycotic infections in Sudan

The most prevalent fungal infection in Sudan based on our estimates is tinea capitis, a superficial disease caused by dermatophytes and primarily affects children of school age. Dermatophytoses are the most common fungal infections worldwide infecting around 20%-25% of the world’s population [80]. In Africa, around 138.1 million children suffer from tinea capitis, which is around one out of every five children [75]. Thus, our estimates of 4,127,760 case in Sudan is not surprising and is close to what has been reported in other Sub-Saharan countries [75]. Furthermore, tinea capitis is highly contagious infection; of the 117 patients studied by Magzoub et al. and attended Khartoum Dermatological Hospital, 82.9% reported a history of contact with infected patients [81]. Unlike skin and nail dermatophytosis, scalp infection is a morbid condition that requires prolonged antifungal medication and might cause inflammation resulting in permanent hair loss. The British Association of Dermatologists recommends the use of terbinafine for treating infections by *Trichophyton* and griseofulvin for treating cases of *Microsporum* spp. [82]. Therefore, optimal diagnosis should not only be made by direct microscopy, but also by culture to allow species identification. In Sudan, direct microscopy is performed occasionally in specialized laboratories and culture is rarely performed [83] (Table 3).

**Table 3:**
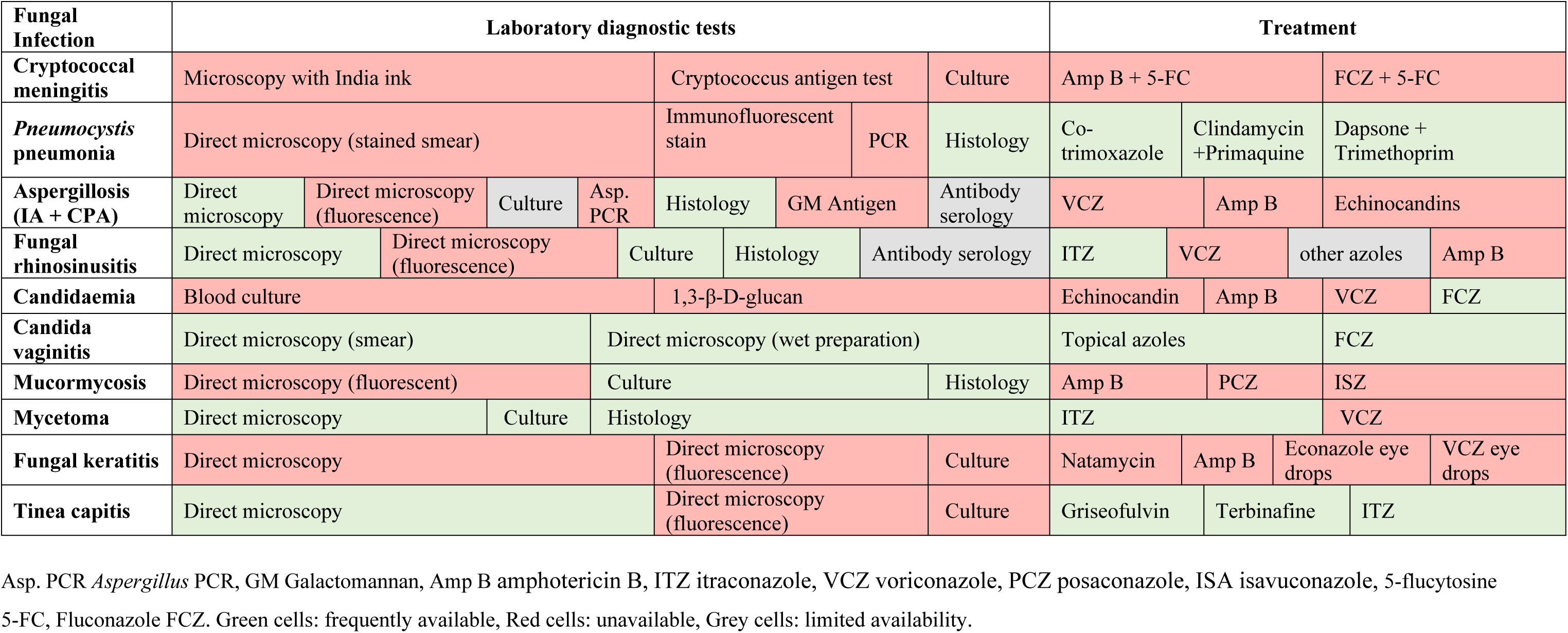
Availability of laboratory diagnostic methods and antifungal medications for fungal infections in Sudan [90]

Fungal keratitis is a potentially blinding condition that annually affects over 1 million individuals from all over the world [84]. The burden is however higher in tropical and subtropical areas; in a study from East Africa, fungal keratitis accounted for 25% of the total microbial keratitis, and in Egypt it was 55% [85, 86]. Of the 64,529 Sudanese patients studied by Lakho et al. [87], 1279 (3.8%) had an eye infection, but the type of microorganism was not determined. To estimate the burden in Sudan, we applied the estimates from Egypt (14 per 100,000) [76]. Mycotic keratitis is considered an ophthalmic emergency requiring rapid intervention to prevent ocular morbidity and eye loss. Detection of fungal elements in the corneal scraping followed by culture are the gold standard diagnostic procedures [84]. In Sudan, fungal culture is rarely performed, and direct microscopy of corneal scraping is only available in limited situations (Table 2). Fungal culture is needed to identify the etiology: more than 100 species of fungi are reported to cause fungal keratitis, among them is the multidrug resistant *Fusarium* spp. [84]. Natamycin eye drops, a drug that was added to the WHO list of essential medicines in 2017 is the first line antifungal therapy for treating fungal keratitis [88]. Nevertheless, based on GAFFI’s surveys, this drug is not available in many countries in the world including Sudan [89]. Most of the cases estimated in our study probably have gone undiagnosed or not received proper treatment. With the current lack of diagnosis and treatment of fungal keratitis globally, scientists have called for the disease to be added to the WHO list of neglected tropical diseases [84].

Sudan is the country where the highest number of mycetoma cases are present; the country is described as the disease homeland. The first report of mycetoma in Sudan is back to the 18^th^ century, yet diagnosis and treatment remain challenging. In 1991, the MRC was established in Khartoum to provide medical care for the patients and to promote research [17]. After the recognition of mycetoma as a neglected tropical disease, the MRC has officially become a WHO Collaborating Center. Recently, researchers from the center have tried to estimate the burden of the disease for a period of 28 years; the overall number of people who have been suffering from mycetoma was 63,825 cases of which eumycetoma accounted for 51,541 (81%) cases [71]. However, comprehensive epidemiologic data on mortality or DALY expectancy related to this disabling condition is currently unavailable. Eumycetoma prevalence based on our study was 36 per 100,000. This rate is remarkably high, and the burden of other serious fungal diseases estimated in the present study are high too. For example, pulmonary aspergillosis (CPA and IA) affects 18,338 individuals, and fungal keratitis affect 6,552; both infections require extensive diagnostic procedures and antifungal therapy but are not on the list of the WHO of neglected diseases. Thus, not only mycetoma, but other serious fungal diseases are neglected too.

Although our estimates showed a high burden of fungal infections in immunocompetent individuals, we also have found a high burden in patients with underlying disorders that are more susceptible or at risk for opportunistic fungal diseases. Among the most affected population are HIV/AIDS and TB patients. In Sudan, the number of HIV cases is relatively low, but tuberculosis is remarkably high. In 2020, around 28,000 Sudanese were infected by *M. tuberculosis*, and this number will probably increase as a result of current conflict and economic instability [91]. Survivors of TB may develop CPA in 6.5% of individuals with remaining cavities, each year, and a significant minority of those thought to have pulmonary TB with negative tests probably have CPA [43]. Globally, CPA is thought to affect around 3 million individuals of whom 1.2 million are post-TB [44]. In Sudan, despite the large number of TB infected individuals, CPA is rarely diagnosed. In 1972, Mahgoub and Elhassan reported a case of pulmonary aspergillosis with cavitation, probably representing CPA [92]. Furthermore, *Aspergillus* spp. was isolated from 9 out of 150 TB patients studied by Baraka et al. in Khartoum [93]. According to our estimates around 3,000 TB survivors are affected by CPA annually. This is similar to what has been reported in Ethiopia, a country with a high burden of TB like Sudan [14]. CPA is associated with a high mortality rate; estimated to be 1,338 annually in the current study (data not shown). Thus, patients with persistent pulmonary symptoms but smear negative should be screened for CPA; presence of cavities in X-ray combined with serology or direct detection and isolation of *Aspergillus* from respiratory samples are the main diagnostic criteria [94]. Surprisingly, the above-mentioned tests might be available in Sudan, but the detection rate of CPA patients is extremely low. That is likely due to the inadequate awareness of physicians and healthcare professionals. During the period of 4 years, the mycology reference laboratory in Khartoum received only 713 sputum samples from patients with suspected fungal infections. Underlying diseases in those patients were not only TB, but also other respiratory conditions such as asthma [95].

Fungal allergy (sensitization) and airways infection affect asthmatic patients and are usually manifest as SAFS and ABPA (otherwise known as ‘fungal asthma’), with some overlap between the two conditions. Fungal asthma is usually linked to poorly controlled asthma and responsive to antifungal therapy [96]. Our estimate for SAFS is 86,860 and for ABPA is 67,860, placing them as the fourth most common fungal diseases in Sudan. Compared to other neighboring countries, these estimates are relatively lower than in Egypt, but similar to the Democratic Republic of Congo [76, 97]. In Sudan, asthma is among the top 10 cause of hospitalization, and the rate of uncontrolled disease is very high reaching up to 84.5% according to a recent study from Osman et al. [98].

In general, the prevalence of asthma and other respiratory diseases such as COPD in Africa is high, particularly in the sub-Saharan countries. Among the main risk factors are tobacco smoking and air pollution resulting from the use of unprocessed biomass fuels. Around 90% of rural population in Africa rely on biomass fuels for cooking, and that holds true for Sudan, as half of Sudanese population reside in rural area [99]. The estimated prevalence of COPD in Sudan is 2,214,950 cases of whom 232,570 are probably hospitalized, annually [25]. Those patients are at risk of invasive aspergillosis; we estimated an annual incidence of 3,040 cases of IA in COPD patients. This needs further study. Invasive aspergillosis also affects patients with malignancies, transplantation, and AIDS. With the emergence of COVID-19, another population has become at risk of infection [100]. Invasive infections by *Aspergillus* are generally associated with high morbidity and mortality. Therefore, WHO placed *Aspergillus fumigatus* among the critical group in the priority list of pathogenic fungi [101]. A consensus guideline for the treatment of invasive aspergillosis recommends the use of voriconazole as first line therapy or, if not available, then isavuconazole, posaconazole, amphotericin B or an echinocandin [102]. Unfortunately, all these drugs are not available in Sudan and of all the azoles only itraconazole and fluconazole are available [89] (Table 3).

According to UNAIDS, deaths from AIDS-related illness in Sudan in 2021 were 2,300, of which fungal infections are probably among the major causes. Every year, around 135,000 people in sub-Saharan Africa lose their life because of cryptococcal meningitis [8]. We estimated the annual number of new cases of CM and PCP among adult HIV/AIDS in Sudan to 462 and 571, respectively. These estimates are considerably low compared to other countries in Africa [8, 14]. We did not estimate the burden in non-HIV populations. Nevertheless, we could not find a single report from Sudan for the two diseases in both HIV and non-HIV individuals. That is probably due to the lack of diagnostic methods in the country. Lumber puncture is rarely performed and cryptococcal capsular antigen (CrAg) test is not available either (Table 3). Detection of *P. jirovecii* in induced sputum or bronchoalveolar lavage from suspected cases is the gold standard procedure for the diagnosis of PCP [103]. Microscopy is simple without requiring sophisticated instruments although *Pneumocystis* PCR is more sensitive [103]. However, the lack of awareness and training among the physicians and laboratory personnel is probably the main reasons for under-reporting and under recognition of cases. Of note that CM, PCP, and histoplasmosis are increasingly reported in Africa [104, 105]. A recent study from Nigeria found that 7.7% of HIV patients with low CD4 counts tested positive for *Histoplasma* antigen [104]. In Sudan, there are probably more cases of *Histoplasma* than the two cases of African histoplasmosis reported in 1988 and 2008 [37, 38].

*Candida* and *Candida*-like yeasts are among the most common cause of fungal infections globally. Invasive candidiasis is associated with high economic burden and high mortality rate - up to 50% [58]. Based on our estimates, *Candida* related infections are the second most common fungal infections in Sudan. Although mostly mucocutaneous, we estimated candidemia in around 2300 individual every year. The recommended antifungal therapy for candidemia in neutropenic and critically ill patients is echinocandins which is unavailable in Sudan, but fluconazole is available (Table 3) [89]. Prior to the antifungal therapy, identification of the yeast down to the species level is paramount especially after the emergence of the multi-drug resistant *C. auris*, a species causing outbreaks in many countries of the world and has been reported from Sudan [106].

In general, for most of the mycotic diseases that are probably present in Sudan, diagnostic methods are lacking, and this could explain the scarcity of published data on these infections (Table 3). Nevertheless, we were able to estimate the burden using data and models utilized to study fungal diseases in other countries. Indeed, our estimates might not reflect the real situation, but they give some perspective on a high burden of some fungal diseases in the country. For example, it is unlikely that there are no cases for cryptococcosis, PCP or pulmonary aspergillosis are present in the Sudan, given with the substantial number of asthmatic, TB and HIV patients. Thus, obviously fungal diseases are badly neglected and require attention by healthcare authorities. There is an urgent need for campaigns to increase the awareness, promote research, and develop national surveillance program. Healthcare and academic institutions should take initiative by developing research and provide training for medical staff. Of note, there is a mycology reference laboratory in Khartoum located at the National Public Health Laboratory and run by the Faculty of Medicine, University of Khartoum. Such laboratories must be sufficiently well equipped to meet the requirement for diagnosis of fungal diseases.

In conclusion, the burden of serious fungal infections in Sudan has been estimated. Remarkably, of every 100,000 Sudanese women 2,698 suffer from recurrent *Candida* vaginitis. In addition, the 6,552 patients suffering from fungal keratitis are probably at risk of blindness in the absence of proper diagnosis and antifungal therapy. Awareness of morbidity and mortality of mycotic diseases need to be addressed urgently. Furthermore, local epidemiologic data is needed to validate the estimated burden and to provide more precise figures for the diseases in the country.

## Data Availability

All data produced in the present work are contained in the manuscript

## References

1. Coates MM, Ezzati M, Robles Aguilar G, Kwan GF, Vigo D, Mocumbi AO, et al. Burden of disease among the world’s poorest billion people: An expert-informed secondary analysis of Global Burden of Disease estimates. PLoS One. 2021;16(8):e0253073. Epub 20210816. doi: 10.1371/journal.pone.0253073. PubMed PMID: 34398896; PubMed Central PMCID: PMCPMC8366975.

2. World Health Organization (WHO). WHO methods and data sources for country-level causes of death 2000-2019 2020 [cited 2023 01-03-2023]. Available from: https://cdn.who.int/media/docs/default-source/gho-documents/global-health-estimates/ghe2019_cod_methods.pdf.

3. Boutayeb A. The Impact of Infectious Diseases on the Development of Africa. Handbook of Disease Burdens and Quality of Life Measures. 2010:1171–88.

4. Kharsany AB, Karim QA. HIV Infection and AIDS in Sub-Saharan Africa: Current Status, Challenges and Opportunities. Open AIDS J. 2016;10:34–48. Epub 20160408. doi: 10.2174/1874613601610010034. PubMed PMID: 27347270; PubMed Central PMCID: PMCPMC4893541.

5. Chakaya J, Khan M, Ntoumi F, Aklillu E, Fatima R, Mwaba P, et al. Global Tuberculosis Report 2020 - Reflections on the Global TB burden, treatment and prevention efforts. Int J Infect Dis. 2021;113 Suppl 1(Suppl 1):S7–S12. Epub 20210311. doi: 10.1016/j.ijid.2021.02.107. PubMed PMID: 33716195; PubMed Central PMCID: PMCPMC8433257.

6. Jarvis JN, Boulle A, Loyse A, Bicanic T, Rebe K, Williams A, et al. High ongoing burden of cryptococcal disease in Africa despite antiretroviral roll out. AIDS. 2009;23(9):1182–3. doi: 10.1097/QAD.0b013e32832be0fc. PubMed PMID: 19451796; PubMed Central PMCID: PMCPMC2871312.

7. Oladele R, Ogunsola F, Akanmu A, Stocking K, Denning DW, Govender N. Opportunistic fungal infections in persons living with advanced HIV disease in Lagos, Nigeria; a 12-year retrospective study. Afr Health Sci. 2020;20(4):1573–81. doi: 10.4314/ahs.v20i4.9. PubMed PMID: 34394217; PubMed Central PMCID: PMCPMC8351866.

8. Rajasingham R, Smith RM, Park BJ, Jarvis JN, Govender NP, Chiller TM, et al. Global burden of disease of HIV-associated cryptococcal meningitis: an updated analysis. Lancet Infect Dis. 2017;17(8):873–81. Epub 20170505. doi: 10.1016/S1473-3099(17)30243-8PubMed PMID: 28483415; PubMed Central PMCID: PMCPMC5818156.

9. Global Action for Fungal Infections. GAFFI Roadmap [cited 2023 01-03-2023]. Available from: https://gaffi.org/roadmap/.

10. Bongomin F, Gago S, Oladele RO, Denning DW. Global and Multi-National Prevalence of Fungal Diseases-Estimate Precision. J Fungi (Basel). 2017;3(4). Epub 20171018. doi: 10.3390/jof3040057. PubMed PMID: 29371573; PubMed Central PMCID: PMCPMC5753159.

11. Rodrigues ML, Nosanchuk JD. Fungal diseases as neglected pathogens: A wake-up call to public health officials. PLoS Negl Trop Dis. 2020;14(2):e0007964. Epub 20200220. doi: 10.1371/journal.pntd.0007964. PubMed PMID: 32078635; PubMed Central PMCID: PMCPMC7032689.

12. de Hoog GS GJ, Gené J, Ahmed SA, Al-Hatmi AMS, Figueras MJ, Vitale RG. Atlas of Clinical Fungi. 4 ed. Hilversum: Foundation Atlas of Clinical fungi; 2020.

13. Global Action for Fungal Infections. Country Fungal Disease Burden [cited 2023 01-03-2023]. Available from: https://gaffi.org/media/country-fungal-disease-burdens.

14. Tufa TB, Denning DW. The Burden of Fungal Infections in Ethiopia. J Fungi (Basel). 2019;5(4). Epub 20191122. doi: 10.3390/jof5040109. PubMed PMID: 31771096; PubMed Central PMCID: PMCPMC6958437.

15. Werkneh S, Orefuwa E, Denning DW. Current situation of fungal diseases in Eritrea. Mycoses. 2022;65(8):806–14. Epub 20220619. doi: 10.1111/myc.13474. PubMed PMID: 35633079; PubMed Central PMCID: PMCPMC9545796.

16. United Nations Development Programme (UNDP) [cited 2023 01-03-2023]. Available from: https://www.undp.org/sudan.

17. Fahal A, Mahgoub el S, El Hassan AM, Abdel-Rahman ME. Mycetoma in the Sudan: an update from the Mycetoma Research Centre, University of Khartoum, Sudan. PLoS Negl Trop Dis. 2015;9(3):e0003679. Epub 20150327. doi: 10.1371/journal.pntd.0003679. PubMed PMID: 25816316; PubMed Central PMCID: PMCPMC4376889.

18. Zijlstra EE, van de Sande WW, Fahal AH. Mycetoma: A Long Journey from Neglect. PLoS Negl Trop Dis. 2016;10(1):e0004244. Epub 20160121. doi: 10.1371/journal.pntd.0004244. PubMed PMID: 26797103; PubMed Central PMCID: PMCPMC4721668.

19. The World Factbook - CIA [cited 2023 01-03-2023]. Available from: https://www.cia.gov/the-world-factbook/countries/sudan/.

20. United Nations, Department of Economic and Social Affairs, Population Division. World Population Prospects 2022, Online Edition. 2022 [cited 2023 01-03-2023]. Available from: https://population.un.org/wpp/Download/Standard/MostUsed/.

21. The Joint United Nations Programme on HIV/AIDS (UNAIDS). Sudan Fact Sheet. 2021 [cited 2023 01-03-2023]. Available from: https://www.unaids.org/en/regionscountries/countries/sudan.

22. Denning DW. Minimizing fungal disease deaths will allow the UNAIDS target of reducing annual AIDS deaths below 500 000 by 2020 to be realized. Philos Trans R Soc Lond B Biol Sci. 2016;371(1709). doi: 10.1098/rstb.2015.0468. PubMed PMID: 28080991; PubMed Central PMCID: PMCPMC5095544.

23. World Health Organization (WHO). Global tuberculosis report 2020 [cited 2023 01-03-2023]. Available from: https://www.who.int/publications/i/item/9789240013131.

24. To T, Stanojevic S, Moores G, Gershon AS, Bateman ED, Cruz AA, et al. Global asthma prevalence in adults: findings from the cross-sectional world health survey. BMC Public Health. 2012;12:204. Epub 20120319. doi: 10.1186/1471-2458-12-204. PubMed PMID: 22429515; PubMed Central PMCID: PMCPMC3353191.

25. Egere U, Shayo E, Ntinginya N, Osman R, Noory B, Mpagama S, et al. Management of chronic lung diseases in Sudan and Tanzania: how ready are the country health systems? BMC Health Serv Res. 2021;21(1):734. Epub 20210724. doi: 10.1186/s12913-021-06759-9. PubMed PMID: 34303370; PubMed Central PMCID: PMCPMC8310588.

26. International Agency for Research on Cancer. Global Cancer Observatory. 2020 [cited 2023 01-03-2023]. Available from: https://gco.iarc.fr/today/data/factsheets/populations/729-sudan-fact-sheets.pdf.

27. Union for International Cancer Control. Acute Myelogenous Leukemia and Acute Promyelocytic Leukemia. 2014 Review of Cancer Medicines on the WHO List of Essential Medicines. [cited 2023 01-03-2023]. Available from: https://silo.tips/download/acute-myelogenous-leukemia-including-acute-promyelocytic-leukemia.

28. International Registry in Organ Donation and Transplantation (IRODaT) [cited 2023 01-03-2023]. Available from: https://www.irodat.org/.

29. The World Bank [cited 2023 01-03-2023]. Available from: https://data.worldbank.org/country/SD.

30. Wharton G, Ali OE, Khalil S, Yagoub H, Mossialos E. Rebuilding Sudan’s health system: opportunities and challenges. Lancet. 2020;395(10219):171–3. doi: 10.1016/S0140-6736(19)32974-5. PubMed PMID: 31954447.

31. World Health Organization (WHO). Eastern Mediterranean Region Framework for health information systems and core indicators for monitoring health situation and health system performance 2016 [cited 2023 01-03-2023]. Available from: https://applications.emro.who.int/dsaf/EMROPUB_2016_EN_19169.pdf?ua=1.

32. Tageldin MA, Nafti S, Khan JA, Nejjari C, Beji M, Mahboub B, et al. Distribution of COPD-related symptoms in the Middle East and North Africa: results of the BREATHE study. Respir Med. 2012;106 Suppl 2:S25–32. doi: 10.1016/S0954-6111(12)70012-4. PubMed PMID: 23290701.

33. Polatli M, Bilgin C, Saylan B, Baslilar S, Toprak E, Ergen H, et al. A cross sectional observational study on the influence of chronic obstructive pulmonary disease on activities of daily living: the COPD-Life study. Tuberk Toraks. 2012;60(1):1–12. doi: 10.5578/tt.3414. PubMed PMID: 22554361.

34. Smith E, Orholm M. Trends and patterns of opportunistic diseases in Danish AIDS patients 1980-1990. Scand J Infect Dis. 1990;22(6):665–72. doi: 10.3109/00365549009027119. PubMed PMID: 2135639.

35. Serraino D, Puro V, Boumis E, Angeletti C, Girardi E, Petrosillo N, et al. Epidemiological aspects of major opportunistic infections of the respiratory tract in persons with AIDS: Europe, 1993-2000. AIDS. 2003;17(14):2109–16. doi: 10.1097/00002030-200309260-00012. PubMed PMID: 14502014.

36. Kuate MPN, Ekeng BE, Kwizera R, Mandengue C, Bongomin F. Histoplasmosis overlapping with HIV and tuberculosis in sub-Saharan Africa: challenges and research priorities. Ther Adv Infect Dis. 2021;8:20499361211008675. Epub 20210409. doi: 10.1177/20499361211008675. PubMed PMID: 33889408; PubMed Central PMCID: PMCPMC8040546.

37. Gumaa SA, Ahmed MA, Hassan ME, Hassan AM. A case of African histoplasmosis from Sudan. Trans R Soc Trop Med Hyg. 1988;82(3):503–5. doi: 10.1016/0035-9203(88)90178-2. PubMed PMID: 3232196.

38. Mudawi HM, Elamin EM, Baraka OZ, El-Hassan AM. Addison’s disease due to Histoplasma duboisii infection of the adrenal glands. Saudi Med J. 2008;29(6):904–6. PubMed PMID: 18521476.

39. Lortholary O, Gangneux JP, Sitbon K, Lebeau B, de Monbrison F, Le Strat Y, et al. Epidemiological trends in invasive aspergillosis in France: the SAIF network (2005-2007). Clin Microbiol Infect. 2011;17(12):1882–9. Epub 20110610. doi: 10.1111/j.1469-0691.2011.03548.x. PubMed PMID: 21668573.

40. Chen CY, Sheng WH, Tien FM, Lee PC, Huang SY, Tang JL, et al. Clinical characteristics and treatment outcomes of pulmonary invasive fungal infection among adult patients with hematological malignancy in a medical centre in Taiwan, 2008-2013. J Microbiol Immunol Infect. 2020;53(1):106–14. Epub 20180131. doi: 10.1016/j.jmii.2018.01.002. PubMed PMID: 29449166.

41. Denning DW, Morgan EF. Quantifying Deaths from Aspergillosis in HIV Positive People. J Fungi (Basel). 2022;8(11). Epub 20221027. doi: 10.3390/jof8111131. PubMed PMID: 36354898; PubMed Central PMCID: PMCPMC9693143.

42. Bongomin F. Post-tuberculosis chronic pulmonary aspergillosis: An emerging public health concern. PLoS Pathog. 2020;16(8):e1008742. Epub 20200820. doi: 10.1371/journal.ppat.1008742. PubMed PMID: 32817649; PubMed Central PMCID: PMCPMC7440622.

43. Denning DW, Cole DC, Ray A. New estimation of the prevalence of chronic pulmonary aspergillosis (CPA) related to pulmonary TB - a revised burden for India. IJID Reg. 2023;6:7–14. Epub 20221118. doi: 10.1016/j.ijregi.2022.11.005. PubMed PMID: 36568568; PubMed Central PMCID: PMCPMC9772841.

44. Denning DW, Pleuvry A, Cole DC. Global burden of chronic pulmonary aspergillosis as a sequel to pulmonary tuberculosis. Bull World Health Organ. 2011;89(12):864–72. Epub 20110927. doi: 10.2471/BLT.11.089441. PubMed PMID: 22271943; PubMed Central PMCID: PMCPMC3260898.

45. Musa O MA, Elsony A, Eltigani M, Elmahi G, Elawad A, Dawoud O. Prevalence and Risk Factors of Asthma Symptoms in Adult Sudanese Using a Modified ISAAC. International Journal of Science and Research. 2016;5(2):1153–6.

46. Ait-Khaled N, Odhiambo J, Pearce N, Adjoh KS, Maesano IA, Benhabyles B, et al. Prevalence of symptoms of asthma, rhinitis and eczema in 13- to 14-year-old children in Africa: the International Study of Asthma and Allergies in Childhood Phase III. Allergy. 2007;62(3):247–58. doi: 10.1111/j.1398-9995.2007.01325.x. PubMed PMID: 17298341.

47. Binegdie AB, Meme H, El Sony A, Haile T, Osman R, Miheso B, et al. Chronic respiratory disease in adult outpatients in three African countries: a cross-sectional study. Int J Tuberc Lung Dis. 2022;26(1):18–25. doi: 10.5588/ijtld.21.0362. PubMed PMID: 34969424; PubMed Central PMCID: PMCPMC8734192.

48. Denning DW, Pleuvry A, Cole DC. Global burden of allergic bronchopulmonary aspergillosis with asthma and its complication chronic pulmonary aspergillosis in adults. Med Mycol. 2013;51(4):361–70. Epub 20121204. doi: 10.3109/13693786.2012.738312. PubMed PMID: 23210682.

49. Ibrahim SA, Fadl Elmola MA, Karrar ZA, Arabi AM, Abdullah MA, Ali SK, et al. Cystic fibrosis in Sudanese children: First report of 35 cases. Sudan J Paediatr. 2014;14(1):39–44. PubMed PMID: 27493388; PubMed Central PMCID: PMCPMC4949914.

50. Benatar SR, Keen GA, Du Toit Naude W. Aspergillus hypersensitivity in asthmatics in Cape Town. Clin Allergy. 1980;10(3):285–91. doi: 10.1111/j.1365-2222.1980.tb02109.x. PubMed PMID: 7418186.

51. Al-Mobeireek AF, El-Rab M, Al-Hedaithy SS, Alasali K, Al-Majed S, Joharjy I. Allergic bronchopulmonary mycosis in patients with asthma: period prevalence at a university hospital in Saudi Arabia. Respir Med. 2001;95(5):341–7. doi: 10.1053/rmed.2001.1047. PubMed PMID: 11392574.

52. Kwizera R, Bongomin F, Olum R, Meya DB, Worodria W, Bwanga F, et al. Fungal asthma among Ugandan adult asthmatics. Medical Mycology. 2021;59(9):923–33. doi: 10.1093/mmy/myab023.

53. Singh V. Fungal Rhinosinusitis: Unravelling the Disease Spectrum. J Maxillofac Oral Surg. 2019;18(2):164–79. Epub 20190128. doi: 10.1007/s12663-018-01182-w. PubMed PMID: 30996535; PubMed Central PMCID: PMCPMC6441414.

54. Said SA, McHembe MD, Chalya PL, Rambau P, Gilyoma JM. Allergic rhinitis and its associated co-morbidities at Bugando Medical Centre in Northwestern Tanzania; A prospective review of 190 cases. BMC Ear Nose Throat Disord. 2012;12:13. Epub 20121108. doi: 10.1186/1472-6815-12-13. PubMed PMID: 23136895; PubMed Central PMCID: PMCPMC3515478.

55. Veress B, Malik OA, el-Tayeb AA, el-Daoud S, Mahgoub ES, el-Hassan AM. Further observations on the primary paranasal aspergillus granuloma in the Sudan: a morphological study of 46 cases. Am J Trop Med Hyg. 1973;22(6):765–72. doi: 10.4269/ajtmh.1973.22.765. PubMed PMID: 4200814.

56. Ahmed SAO, Mustafa OME. Clinical features of Fungal Rhinosinusitis in Sudan. A study of 440 cases. PAN Arab Journal of Rhinology. 2013;3(1).

57. Mahgoub ES, Ismail MAI and Gabr A.. Fungal Sinusitis: Sudanese Experience. Austin J Otolaryngol. 2016;3(4):1085.

58. Montravers P, Mira JP, Gangneux JP, Leroy O, Lortholary O, AmarCand study g. A multicentre study of antifungal strategies and outcome of Candida spp. peritonitis in intensive-care units. Clin Microbiol Infect. 2011;17(7):1061–7. Epub 20101019. doi: 10.1111/j.1469-0691.2010.03360.x. PubMed PMID: 20825438.

59. Mushi MF, Olum R, Bongomin F. Prevalence, antifungal susceptibility and etiology of vulvovaginal candidiasis in sub-Saharan Africa: a systematic review with meta-analysis and meta-regression. Med Mycol. 2022;60(7). doi: 10.1093/mmy/myac037. PubMed PMID: 35781514.

60. Kafi SK, Mohamed AO, Musa HA. Prevalence of sexually transmitted diseases (STD) among women in a suburban Sudanese community. Ups J Med Sci. 2000;105(3):249–53. doi: 10.3109/2000-1967-179. PubMed PMID: 11261611.

61. Nemery HM. Phenotypic Characterization of Vaginal Candidiasis in Sudanese Pregnant Women African Journal of Medical Sciences. 2019;4(4).

62. Abdelaziz ZA, Ibrahim ME, Bilal NE, Hamid ME. Vaginal infections among pregnant women at Omdurman Maternity Hospital in Khartoum, Sudan. J Infect Dev Ctrie. 2014;8(4):490–7. Epub 20140415. doi: 10.3855/jidc.3197. PubMed PMID: 24727516.

63. Denning DW, Kneale M, Sobel JD, Rautemaa-Richardson R. Global burden of recurrent vulvovaginal candidiasis: a systematic review. Lancet Infect Dis. 2018;18(11):e339–e47. Epub 20180802. doi: 10.1016/S1473-3099(18)30103-8. PubMed PMID: 30078662.

64. Lynch JB, Husband AD. Subcutaneous phycomycetosis. J Clin Pathol. 1962;15(2):126–32. doi: 10.1136/jcp.15.2.126. PubMed PMID: 14467579; PubMed Central PMCID: PMCPMC480359.

65. Fischer N, Ruef C, Ebnother C, Bachli EB. Rhinofacial Conidiobolus coronatus infection presenting with nasal enlargement. Infection. 2008;36(6):594–6. Epub 20081108. doi: 10.1007/s15010-008-8056-5. PubMed PMID: 18998052.

66. Mohammed SA, Abdelsatir AA, Abdellatif M, Suliman SH, Elbasheer OMI, Abdalla AR, et al. Challenging Presentations of Seven Cases of Gastrointestinal Basidiobolomycosis in Sudan: Clinical Features, Histology, Imaging, and Recommendations. J Lab Physicians. 2020;12(4):281–4. Epub 20201230. doi: 10.1055/s-0040-1721149. PubMed PMID: 33390679; PubMed Central PMCID: PMCPMC7773445.

67. Taha S MA, Hassan L, EL Hassan AM, Mustafa G, Yaseen DE. Gastrointestinal Basidiobolomycosis Mimicking Colon Cancer in a Sudanese Patient. 2011;6(1). doi: 10.4314/sjms.v6i1.67277.

68. Gangneux JP, Bougnoux ME, Hennequin C, Godet C, Chandenier J, Denning DW, et al. An estimation of burden of serious fungal infections in France. J Mycol Med. 2016;26(4):385–90. Epub 20161122. doi: 10.1016/j.mycmed.2016.11.001. PubMed PMID: 27887809.

69. Fahal A, Mahgoub el S, El Hassan AM, Abdel-Rahman ME, Alshambaty Y, Hashim A, et al. A new model for management of mycetoma in the Sudan. PLoS Negl Trop Dis. 2014;8(10):e3271. Epub 20141030. doi: 10.1371/journal.pntd.0003271. PubMed PMID: 25356640; PubMed Central PMCID: PMCPMC4214669.

70. Abbott P. Mycetoma in the Sudan. Trans R Soc Trop Med Hyg. 1956;50(1):11–24. doi: 10.1016/0035-9203(56)90004-9. PubMed PMID: 13299347.

71. Hassan R, Cano J, Fronterre C, Bakhiet S, Fahal A, Deribe K, et al. Estimating the burden of mycetoma in Sudan for the period 1991-2018 using a model-based geostatistical approach. PLoS Negl Trop Dis. 2022;16(10):e0010795. Epub 20221014. doi: 10.1371/journal.pntd.0010795. PubMed PMID: 36240229; PubMed Central PMCID: PMCPMC9604875.

72. Gumaa SA. Sporotrichosis in Sudan. Trans R Soc Trop Med Hyg. 1978;72(6):637–40. doi: 10.1016/0035-9203(78)90020-2. PubMed PMID: 734721.

73. Mahgoub ES. Ringworm infection among Sudanese school children. Trans R Soc Trop Med Hyg. 1968;62(2):263–8. doi: 10.1016/0035-9203(68)90167-3. PubMed PMID: 4230638.

74. Ahmed ASAA, Ahmed MAI, Abdelrahma NAM, Adam NKA, Abdel-aziz DS, Ahmed ABM. Prevalence of tinea capitis among school age children in eastern Sudan J Bacteriol Mycol Open Access. 2021;9(2):94‒7. doi: DOI: 10.15406/jbmoa.2021.09.00303.

75. Bongomin F, Olum R, Nsenga L, Namusobya M, Russell L, de Sousa E, et al. Estimation of the burden of tinea capitis among children in Africa. Mycoses. 2021;64(4):349–63. Epub 20201209. doi: 10.1111/myc.13221. PubMed PMID: 33251631.

76. Zaki SM, Denning DW. Serious fungal infections in Egypt. Eur J Clin Microbiol Infect Dis. 2017;36(6):971–4. Epub 20170217. doi: 10.1007/s10096-017-2929-4. PubMed PMID: 28213689.

77. Xu H, Li L, Huang WJ, Wang LX, Li WF, Yuan WF. Invasive pulmonary aspergillosis in patients with chronic obstructive pulmonary disease: a case control study from China. Clin Microbiol Infect. 2012;18(4):403–8. Epub 20111025. doi: 10.1111/j.1469-0691.2011.03503.x. PubMed PMID: 22023558.

78. Matee MI, Scheutz F, Moshy J. Occurrence of oral lesions in relation to clinical and immunological status among HIV-infected adult Tanzanians. Oral Dis. 2000;6(2):106–11. doi: 10.1111/j.1601-0825.2000.tb00110.x. PubMed PMID: 10702788.

79. Buchacz K, Baker RK, Palella FJ, Jr., Chmiel JS, Lichtenstein KA, Novak RM, et al. AIDS-defining opportunistic illnesses in US patients, 1994-2007: a cohort study. AIDS. 2010;24(10):1549–59. doi: 10.1097/QAD.0b013e32833a3967. PubMed PMID: 20502317.

80. Havlickova B, Czaika VA, Friedrich M. Epidemiological trends in skin mycoses worldwide. Mycoses. 2008;51 Suppl 4:2–15. doi: 10.1111/j.1439-0507.2008.01606.x. PubMed PMID: 18783559.

81. Magzoub OS. Tinea Capitis: Frequency and Clinical Manifestations among Children in Khartoum Dermatological Hospital, Sudan. Merit Research Journal of Medicine and Medical Sciences. 2022;10(1):1–8. doi: https://doi.org/10.5281/zenodo.5910631.

82. Fuller LC, Barton RC, Mohd Mustapa MF, Proudfoot LE, Punjabi SP, Higgins EM. British Association of Dermatologists’ guidelines for the management of tinea capitis 2014. Br J Dermatol. 2014;171(3):454–63. doi: 10.1111/bjd.13196. PubMed PMID: 25234064.

83. Badiane AS, Ramarozatovo LS, Doumbo SN, Dorkenoo AM, Mandengue C, Dunaiski CM, et al. Diagnostic capacity for cutaneous fungal diseases in the african continent. Int. J. Dermatol. 2023. (Accepted)

84. Brown L, Kamwiziku G, Oladele RO, Burton MJ, Prajna NV, Leitman TM, et al. The Case for Fungal Keratitis to Be Accepted as a Neglected Tropical Disease. J Fungi (Basel). 2022;8(10). Epub 20221005. doi: 10.3390/jof8101047. PubMed PMID: 36294612; PubMed Central PMCID: PMCPMC9605065.

85. Burton MJ, Pithuwa J, Okello E, Afwamba I, Onyango JJ, Oates F, et al. Microbial keratitis in East Africa: why are the outcomes so poor? Ophthalmic Epidemiol. 2011;18(4):158–63. doi: 10.3109/09286586.2011.595041. PubMed PMID: 21780874; PubMed Central PMCID: PMCPMC3670402.

86. EL Shabrawy RM, El Badawy NEL, Harb AW. The incidence of fungal keratitis in Zagazig University Hospitals, Egypt and the value of direct microscopy and PCR technique in rapid diagnosis. 2013; 3 (4): doi: 105799/ahinjs022013040106. 2013;3(4):186-91. doi: 10.5799/ahinjs.02.2013.04.0106.

87. lakho KA, Mohamed Ali AB. Pattern of eye diseases at tertiary eye hospital in Sudan (Makah Eye Hospital, Khartoum). Albasar Int J Ophthalmol. 2015;3:15–158.

88. Qiu S, Zhao GQ, Lin J, Wang X, Hu LT, Du ZD, et al. Natamycin in the treatment of fungal keratitis: a systematic review and Meta-analysis. Int J Ophthalmol. 2015;8(3):597–602. Epub 20150618. doi: 10.3980/j.issn.2222-3959.2015.03.29. PubMed PMID: 26086015; PubMed Central PMCID: PMCPMC4458670.

89. Global Action For Fungal Infections. Antifungal Drug Maps [cited 2023 01-03-2023]. Available from: https://gaffi.org/antifungal-drug-maps/.

90. Orefuwa E, Gangneux JP, Denning DW. The challenge of access to refined fungal diagnosis: An investment case for low- and middle-income countries. J Mycol Med. 2021;31(2):101140. Epub 20210420. doi: 10.1016/j.mycmed.2021.101140. PubMed PMID: 33971531.

91. Hassanain SA, Edwards JK, Venables E, Ali E, Adam K, Hussien H, et al. Conflict and tuberculosis in Sudan: a 10-year review of the National Tuberculosis Programme, 2004-2014. Confl Health. 2018;12:18. Epub 20180516. doi: 10.1186/s13031-018-0154-0. PubMed PMID: 29785203; PubMed Central PMCID: PMCPMC5954449.

92. Mahgoub ES, el-Hassan AM. Pulmonary aspergillosis caused by Aspergillus flavus. Thorax. 1972;27(1):33–7. doi: 10.1136/thx.27.1.33. PubMed PMID: 4622807; PubMed Central PMCID: PMCPMC472456.

93. Baraka AA, Alabid SA, Mohammed MA, Ahmed NM. Frequency of Pulmonary Aspergillosis Among Clinically Suspected and Under Treatment Tuberculosis Patients, Khartoum State, Sudan. Al-Kindy College Medical Journal. 2022;17(3):180–4. doi: https://doi.org/10.47723/kcmj.v17i3.430.

94. Denning DW. Diagnosing pulmonary aspergillosis is much easier than it used to be: a new diagnostic landscape. Int J Tuberc Lung Dis. 2021;25(7):525–36. doi: 10.5588/ijtld.21.0053. PubMed PMID: 34183097.

95. Ismail M, Gabr A, Zhou S, Mahgoub E, Ahmed S. P346 Pulmonary fungal infection in Sudan, a retrospective study from the Mycology Reference Laboratory. Medical Mycology. 2022;60(Supplement_1). doi: 10.1093/mmy/myac072.P346.

96. Rapeport WG, Ito K, Denning DW. The role of antifungals in the management of patients with severe asthma. Clinical and Translational Allergy. 2020;10(1):46. doi: 10.1186/s13601-020-00353-8.

97. Kamwiziku GK, Makangara JC, Orefuwa E, Denning DW. Serious fungal diseases in Democratic Republic of Congo - Incidence and prevalence estimates. Mycoses. 2021;64(10):1159–69. Epub 20210625. doi: 10.1111/myc.13339. PubMed PMID: 34133799.

98. Osman R AK, ElSony A. Factors associated with uncontrolled asthma among Sudanese adult patients. J Pan Afr Thorac Soc. 2021;2(2):85–93. doi: DOI: 10.25259/JPATS_22_2020.

99. van Gemert F, van der Molen T, Jones R, Chavannes N. The impact of asthma and COPD in sub-Saharan Africa. Prim Care Respir J. 2011;20(3):240–8. doi: 10.4104/pcrj.2011.00027. PubMed PMID: 21509418; PubMed Central PMCID: PMCPMC6549843.

100. Verweij PE, Bruggemann RJM, Azoulay E, Bassetti M, Blot S, Buil JB, et al. Taskforce report on the diagnosis and clinical management of COVID-19 associated pulmonary aspergillosis. Intensive Care Med. 2021;47(8):819–34. Epub 20210623. doi: 10.1007/s00134-021-06449-4. PubMed PMID: 34160631; PubMed Central PMCID: PMCPMC8220883.

101. World Health Organization. WHO fungal priority pathogens list to guide research, development and public health action 2022 [cited 2023 01-03-2023]. Available from: https://www.who.int/publications/i/item/9789240060241.

102. Douglas AP, Smibert OC, Bajel A, Halliday CL, Lavee O, McMullan B, et al. Consensus guidelines for the diagnosis and management of invasive aspergillosis, 2021. Intern Med J. 2021;51 Suppl 7:143–76. doi: 10.1111/imj.15591. PubMed PMID: 34937136.

103. Bateman M, Oladele R, Kolls JK. Diagnosing Pneumocystis jirovecii pneumonia: A review of current methods and novel approaches. Med Mycol. 2020;58(8):1015–28. doi: 10.1093/mmy/myaa024. PubMed PMID: 32400869; PubMed Central PMCID: PMCPMC7657095.

104. Oladele RO, Osaigbovo, II, Akanmu AS, Adekanmbi OA, Ekeng BE, Mohammed Y, et al. Prevalence of Histoplasmosis among Persons with Advanced HIV Disease, Nigeria. Emerg Infect Dis. 2022;28(11):2261–9. doi: 10.3201/eid2811.220542. PubMed PMID: 36286009; PubMed Central PMCID: PMCPMC9622240.

105. Bongomin F, Ekeng BE, Kibone W, Nsenga L, Olum R, Itam-Eyo A, et al. Invasive Fungal Diseases in Africa: A Critical Literature Review. J Fungi (Basel). 2022;8(12). Epub 20221122. doi: 10.3390/jof8121236. PubMed PMID: 36547569; PubMed Central PMCID: PMCPMC9853333.

106. Badri AM, Sherfi SA. First Detection of Emergent Fungal Pathogen Candida auris in Khartoum State, Sudan. J Biomed Sci & Res. 2019;(1). doi: DOI:10.34297/AJBSR.2019.06.000982.

